# Proposing a novel Seriously Deteriorated Patient Indicator (SDPI) for hospitalised ward patients

**DOI:** 10.1101/2025.07.09.25331163

**Authors:** Anton H. van der Vegt, Victoria Campbell, Imogen Mitchell, James Malycha, Ian A. Scott, Arthas Flabouris, Naitik Mehta, Rudolf J. Schnetler, Christopher R Andersen, Daryl Jones

## Abstract

**Objective:** Current deteriorated patient outcome measures, death and unplanned ICU admission (UPICU), don’t consider patients who deteriorate and recover on the ward, nor correctly identify the time that significant deterioration occurs. This limits fair comparative evaluation of Early Warning Tool (EWT) performance and may degrade AI deterioration prediction algorithm accuracy. A Seriously Deteriorated Patient Indicator (SDPI) is required to overcome these limitations.

**Materials and methods:** Using a multi-hospital, retrospective dataset and supported by a clinician committee, we developed the SDPI by (i) identifying a well-embedded baseline EWT with superior identification of patients who would transfer to the ICU in the setting of severe illness, (ii) testing additions/variations in scoring elements to improve that tools accuracy, and (iii) selecting an SDPI threshold above which patients are labelled as seriously deteriorated.

**Results:** 957,445 ward episodes were included (UPICU prevalence 0.4%). The superior baseline tool (AUPRC 0.0752), was successfully augmented by 13.1% (AUPRC 0.085) through 11 adjustments. The final SDPI identified 12,323 seriously deteriorated patients (1.3% of the cohort), of which 8,701 (0.9% of cohort) recovered on the ward, identifying deteriorating patients 7.2 and 71 hours earlier than UPICU or death outcomes respectively.

**Discussion:** This physiologically derived, reproducible SDPI identifies deteriorated patients significantly earlier than UPICU or death and includes a significant cohort of seriously deteriorated patients who would have previously been mislabelled as ‘not deteriorated’.

**Conclusion:** We propose this novel SDPI as part of a composite outcome measure for fairer evaluation of EWT performance and better training of AI prediction models.

## Introduction

Early recognition and management of clinical deterioration in hospitalised patients saves lives.^1,2^ With the rise of Electronic Medical Record (EMR) adoption has come the capability to recognise deterioration based upon Early Warning Tools (EWTs) comprising automated rule-based tools, such as the National Early Warning Score (NEWS),^3^ and higher precision artificial intelligence deterioration prediction algorithms (AI-DPAs)^4^. However, a recent meta-study by Herasevich et al. of the impact of implemented electronic EWTs on hospital mortality and length of stay showed mixed results, with randomised controlled studies showing no improvement.^5^ The included studies used many different outcome definitions, such as cardiopulmonary arrest, death or unplanned admission to the intensive care unit (UPICU). This outcome heterogeneity is a long-standing methodological limitation in the early warning system literature, with Bhavani calling for a consensus on an outcome definition based on, ‘objective physiologic or biological measurements’ with a focus on defining ‘preventable deterioration’.^6^

Although deterioration is broadly defined as ‘a state of worsening’,^7^ there is currently no specific test or physiological specification that defines a deteriorating patient, relying instead on practical surrogates, such as those previously mentioned, that usually signal the end result of deterioration rather than the process of deterioration itself.^4^ These surrogate outcomes have served two important roles: (i) to enable evaluation of the ability of EWTs and AI-DPAs to discriminate between deteriorating and non-deteriorating patients prior to the outcome event; and (ii) to provide training labels (i.e., deterioration event identifiers) that enable AI-DPAs to learn to identify deteriorating patients as early as possible.

However, these commonly used surrogate deterioration events are suboptimal for both purposes, as summarised in **Supplementary Table A1**. A major limitation of the most commonly used outcome events of death and UPICU ^8^ is that many patients who seriously deteriorate on the ward are treated and recover without dying or going to the ICU (**Figure 1, Scenario 2**).^9,10^ These patients will be mislabelled as ‘no deterioration’ and for this sub-cohort, EWT evaluation will be incorrect and AI-DPA training data misleading. Additionally, deterioration often occurs hours or days prior to death or UPICU (**Figure 1, Scenario 4**), meaning that the recorded timepoint of deterioration is incorrect. When using these as deterioration outcome measures, EWTs can claim to ‘predict’ deterioration, when in fact deterioration has already occurred and possibly been acted upon. Also, use of the data captured after deterioration may distort AI-DPA training as it may reflect patients receiving increased monitoring and interventions, or if dying, having intrusive tests, monitoring or interventions withheld. Scenario 3 (**Figure 1**) is valid for both evaluation and AI-DPA training, however, it’s rare in modern hospitals with mature early warning systems.

**Figure 1:**
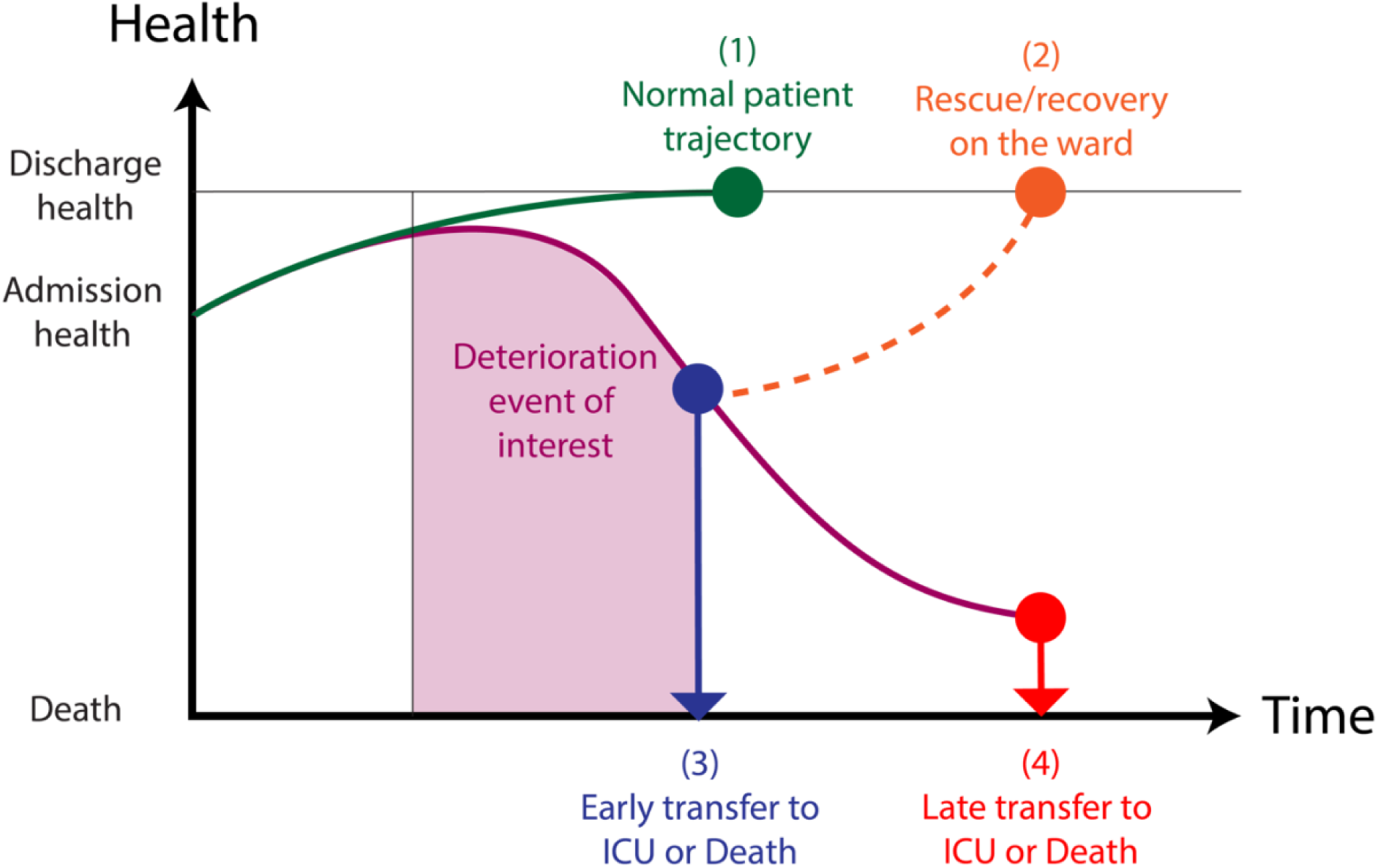
Patient trajectories with respect to deterioration: (1) Most patients will be admitted to the ward, receive appropriate interventions and improve to discharge level of health without deterioration (green); (2) Some patients will have a deterioration event of interest, then recover on the ward (orange). These patients are currently not captured using the existing outcome measures for patient deterioration; (3) Some patients will have a deterioration event and then have an early unplanned admission to ICU or die (Blue). These patients are correctly identified with current outcome measures and the timepoint is also accurate; and (4) Some patients will have a deterioration event of interest, remain on the ward but continue to deteriorate and then have a late admission to ICU or die (Red). These patients are correctly identified using current outcome measures but the timepoint that recorded is inaccurate.

Many of the EWT evaluation and AI-DPA training issues identified above would be addressed by an indicator that more correctly identifies patients who seriously deteriorate (a ‘seriously deteriorated patient’ indicator (SDPI)) and also signals deterioration earlier in its onset, ideally at a time when deterioration is more likely to be preventable or reversible.

### Objective

To develop a novel deterioration outcome measure for seriously deteriorated patients in hospital wards that is physiologically derived from routinely available EMR data and reproducible. We hypothesised that a SDPI outcome would (i) identify deterioration episodes earlier than either death or UPICU; and (ii) identify a large cohort of patients who seriously deteriorate and then recover on the ward, but whose episodes are currently mislabelled as ‘no deterioration’ for EWT evaluation and AI-DPA training.

## Materials and methods

### Study cohort and data collection

Time-stamped vital sign, laboratory and outcome data from all adult ward inpatient admissions were collected between 1 January 2016 and 30 June 2020 at 11 digital public hospitals within Queensland Health, Australia. Sites included four metropolitan and seven regional hospitals. Dataset exclusions are provided in **Supplementary Appendix B**. This study was approved by the Townsville Hospital and Health Service Human Research Ethics Committee (HREC/QTHS/67897) and was granted a waiver of patient consent with Public Health Act approval.

### Stage 1: Selection of a baseline early warning tool for identifying seriously deteriorated patients

Three rule-based EWTs were compared to identify the best performing tool in the range of serious deterioration. Rule-based tools were preferred due to their replicability, explainability and consistency. Two vital sign based tools were selected; (i) NEWS, because it is the most widely used and validated vital sign based scoring tool globally; and (ii) Queensland Adult Deterioration Detection System (Q-ADDS) because it is the most widely used vital sign based scoring tool in Australia, is used with the patients contained in the dataset for this study and has been validated against NEWS.^11^ Laboratory-based Acute Physiology Score Version 2.0 (LAPS2)^12^ was chosen as an implemented, validated rule-based tool which incorporates laboratory results, and has been used for similar purposes in the development of EWTs elsewhere (see all tool details in **Supplementary Appendix C**). These externally validated and clinically implemented tools represented good candidates for a physiologically derived baseline Seriously Deteriorated Patient (SDP) scoring tool to identify seriously deteriorated patients.

#### Evaluation outcome measure

Selection of the best performing of the 3 EWTs was based on how accurately its high score range could identify patients who would transfer to the ICU (UPICU). UPICU was selected over death because the majority of in-hospital deaths in our Australian dataset are not unexpected, similar to the UK and other countries,^13^ and this is increasingly the case in hospitals with mature early warning systems. Also, for reasons previously stated, patients destined to die as expected deaths are not those with the reversible deterioration that we want to recognise. In contrast, a transfer to ICU usually occurs for critically deteriorated patients requiring, or at high risk of requiring, organ support, and who are deemed eligible for such support.

Each admission was divided into ward episodes of care. A single entire admission could have one or more ward episodes, each with either an adverse or non-adverse outcome.^11,12^ For each ward episode, all included data row samples were labelled with either ‘UPICU’, ‘DEATH’ or with ‘NO_OUTCOME’, meaning the patient was discharged alive or converted to a surgical /procedural episode. UPICU was defined as a minimum ICU stay of 6 hours, unless death occurred within that time.^14^

#### Data preparation and baseline early warning tool evaluation

The retrospective dataset was prepared by replacing missing variable values with the last entered value, where it existed. Where a prior value was not entered, each tool applied a zero score (normal) for that variable in its score calculation (see **Supplementary Appendix B3** for missing value counts). Data sample rows were removed when any one or more of heart rate, systolic blood pressure or oxygen saturation (SpO_2_) (the minimum vital sign set provided by standard automated vital sign capture devices) were missing.

Score totals were calculated for all remaining data rows for each tool. A serious deterioration subset of the dataset was then generated for each tool using the following method: (i) The time-stamped data row with the highest score was kept for each episode and all other rows were discarded; and (ii) The highest scoring 5% of episodes were kept and all other episodes discarded. Five percent was selected because 4-5% also aligns with the proportion of patients triggered at the Q-ADDS and NEWS Medical Emergency Team (MET) level at scores 8 and 7, respectively. This process ensured the three EWT-specific datasets were comparable, i.e., containing the same number of episodes and comprising the most seriously deteriorated patient episodes according to each tool.

#### Statistical Analyses

Area Under the Precision-Recall Curve (AUPRC) was used for comparison because unlike Area Under the Receiver Operating Curve (AUROC), it takes into account the highly imbalanced class characteristics of our dataset with UPICU outcome prevalence figures below 1%.^15^ Evaluation was repeated 1000 times to calculate the 95% confidence intervals (CIs) using a bootstrap approach^16^ (random selection with replacement within the tool-specific dataset to the size of the dataset for each bootstrap). If Levene’s test ^17^ for the equality of variances was met, then significant difference between models was calculated using ANOVA with Tukey HSD,^18^ otherwise the non-parametric Mann-Whitney U Test was performed. ^19^ All analyses were performed in Python using sklearn and scipy libraries.

### Experiment Stage 2: Augmenting the baseline SDP scoring tool to improve accuracy

An ablation study was conducted by adding or changing candidate physiological scoring variables (see **Supplementary Table A2**), one at a time, to the baseline tool selected in Stage 1. Dataset preparation, outcome and evaluation methods were the same as those used in Stage 1. Candidate variables, along with their associated scoring ranges where available, were drawn from prior published work and/or clinical rationale, with agreement by clinician committee consisting of 2 ward physicians (VC, IS) and 6 intensivists (DJ, AF, JM, CA, IM, VC (dual specialist)) with an average of 23.5 years of experience and over 190 publications pertaining to patient deterioration and early warning systems.

Individual candidates that resulted in a statistically significant improvement to the AUPRC over the baseline tool were then combined with the baseline tool, to create the final SDP scoring tool.

### Experiment Stage 3: Establishing a Seriously Deteriorated Patient Indicator (SDPI)

To create the final SDPI, a score threshold was required for the SDP scoring tool output from Stage 2, above which patients would be labelled seriously deteriorated. This was achieved through clinical consensus supported by data validating the following criteria: (i) Superior accuracy for identifying UPICU patients than currently implemented MET thresholds for the baseline EWT; (ii) Clinical agreement that the score reflected seriously ill patients; (iii) Similar or earlier timing of recognition of UPICU patients relative to MET alert timing; and (iv) A low chance that the score threshold was reached due to a chronic organ failure or data entry errors. In addition, the odds ratio between each score of the SDP scoring tool and death, UPICU or both were plotted, to provide an orthogonal perspective of the chosen threshold and its practicality.

## Results

### Study Cohort

After exclusions (see Suppl. Appendix B2), there were 831,725 admissions for analysis comprising 957,445 ward episodes involving 343,471 patients and 70,128,956 complete data samples (see Table 1). For transparency, there were 1081 admissions with covid-19 from January – June 2021.

**Table 1:** Dataset characteristics for datasets used in Method stages 1 and 2. Abbreviations, Q-ADDS = Queensland Adult Deterioration Score; NEWS = National Early Warning Score; LAPS-2 = Laboratory-based Acute Physiology Score Version 2.0; IQR = Inter-Quartile Range; ICU = Intensive Care Unit

|  | All episodes dataset | Highest scoring 5% data subset |  |  |
| --- | --- | --- | --- | --- |
|  |  | NEWS scoring dataset | Q-ADDS scoring dataset | LAPS-2 scoring dataset |
| Patients (% of total) | 343,471 | 33,664 (9.8) | 35,182 (10.2) | 30,541 (8.9) |
| Episodes | 957,445 | 47,872 | 47,872 | 47,872 |
| Female percent | 50.2% | 45.1% | 45.6% | 43.1% |
| Median age (IQR) | 59 (40,72) | 69 (56,79) | 68 (53,78) | 73 (61,82) |
| Median hours of episode (IQR) | 13 (3,56) | 122 (58,253) | 119 (47,262) | 127 (62,260) |
| <u>Outcomes: (% of episodes)</u> |  |  |  |  |
| Death | 7,997 (0.8) | 4,985 (10.4) | 4,789 (10.0) | 4,655 (9.7) |
| Unplanned transfer to ICU | 4,074 (0.4) | 1,743 (3.6) | 1,820 (3.8) | 1,421 (3.0) |

### Stage 1: Selection of the baseline Seriously Deteriorated Patient (SDP) scoring tool

Regarding the 3 scoring tools ability to identify patients who would transfer to the ICU, Q-ADDS (AUPRC 0.0752, 0.0698 – 0.0810) was statistically superior, 15.7% higher than NEWS and 22.2% higher than LAPS-2. Differences were significant (see **supplementary Table D1**).

Figure 2 shows the AUPRC for the whole dataset (bottom graph) and then for the tool-specific datasets comprising the highest scoring 5% of patient episodes for each tool (top graph). At lower early warning scores, all tools had similar performance, however at higher scores, the Q-ADDS tool demonstrated superior performance and was consequently selected as the baseline SDP scoring tool.

**Figure 2:**
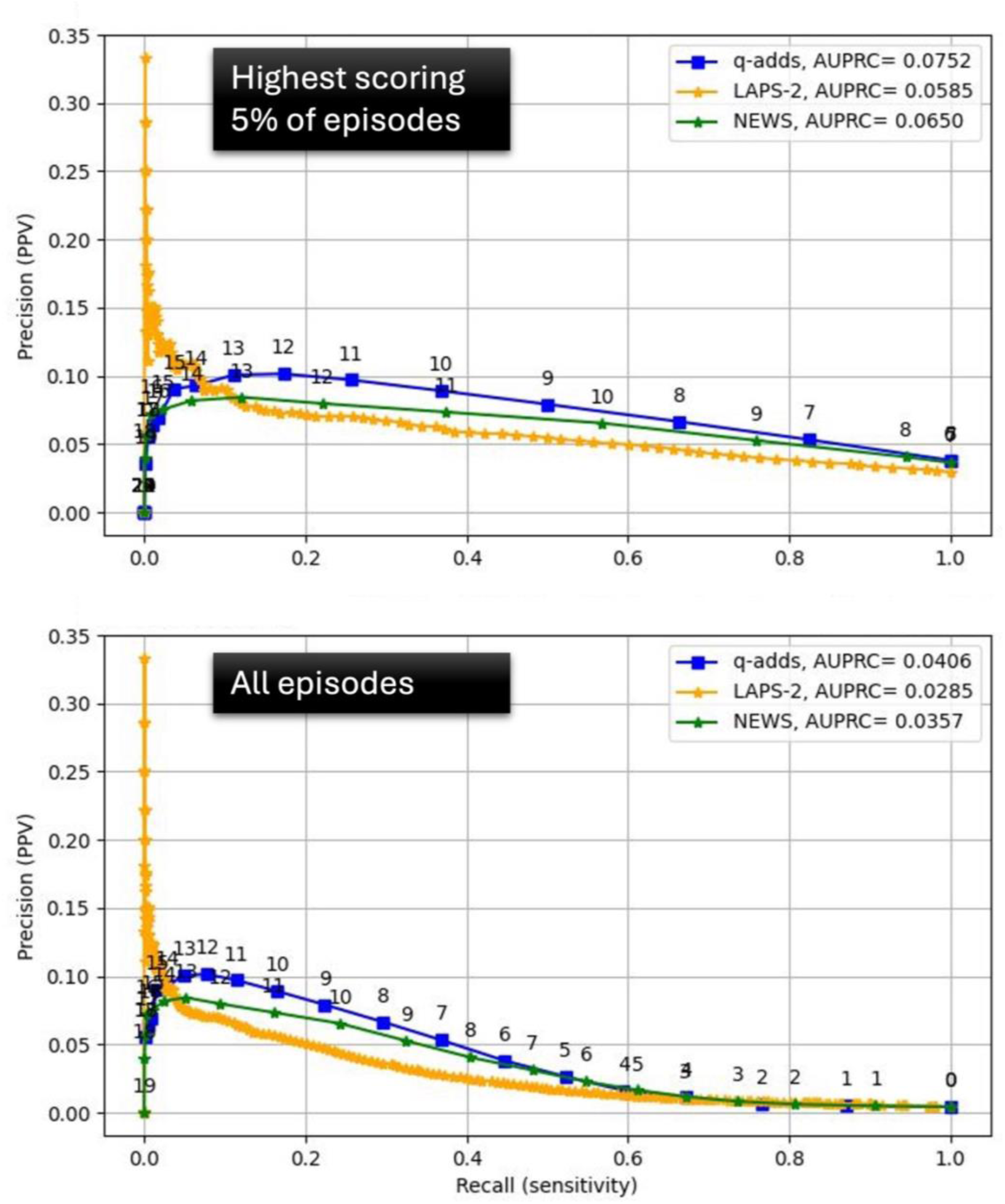
Precision-recall (PR) curves for rule-based index candidates. Top Figure provides PR curves using highest scoring data subset (5% of episodes) for each index. Bottom figure provides PR curves or the all-episode dataset. Abbreviations, PPV = Positive predictive value; q-ADDS = Queensland Adult Deterioration Score; NEWS = National Early Warning Score; LAPS-2 = Laboratory-based Acute Physiology Score Version 2.0; AUPRC = Area under the precision recall curve.

### Stages 2: Augmenting the baseline Seriously Deteriorated Patient (SDP) scoring tool

**Table 2** shows the statistical evaluations of the baseline tool performance after incorporating the new or modified candidate variables to discriminate between patients transferred or not transferred to the ICU. Half of the candidates resulted in a significant improvement with lactate (+6.19%, p-value<0.001) and albumin (+4.97%, p-value<0.001) having the greatest positive impacts. The final augmented SDP scoring tool AUPRC was 13.1% superior (p-value<0.001) to that of the Q-ADDS baseline (**see also supplementary Figure D1**). **Table 3** lists the SDP scoring tool variables and their scoring ranges.

**Table 2:**
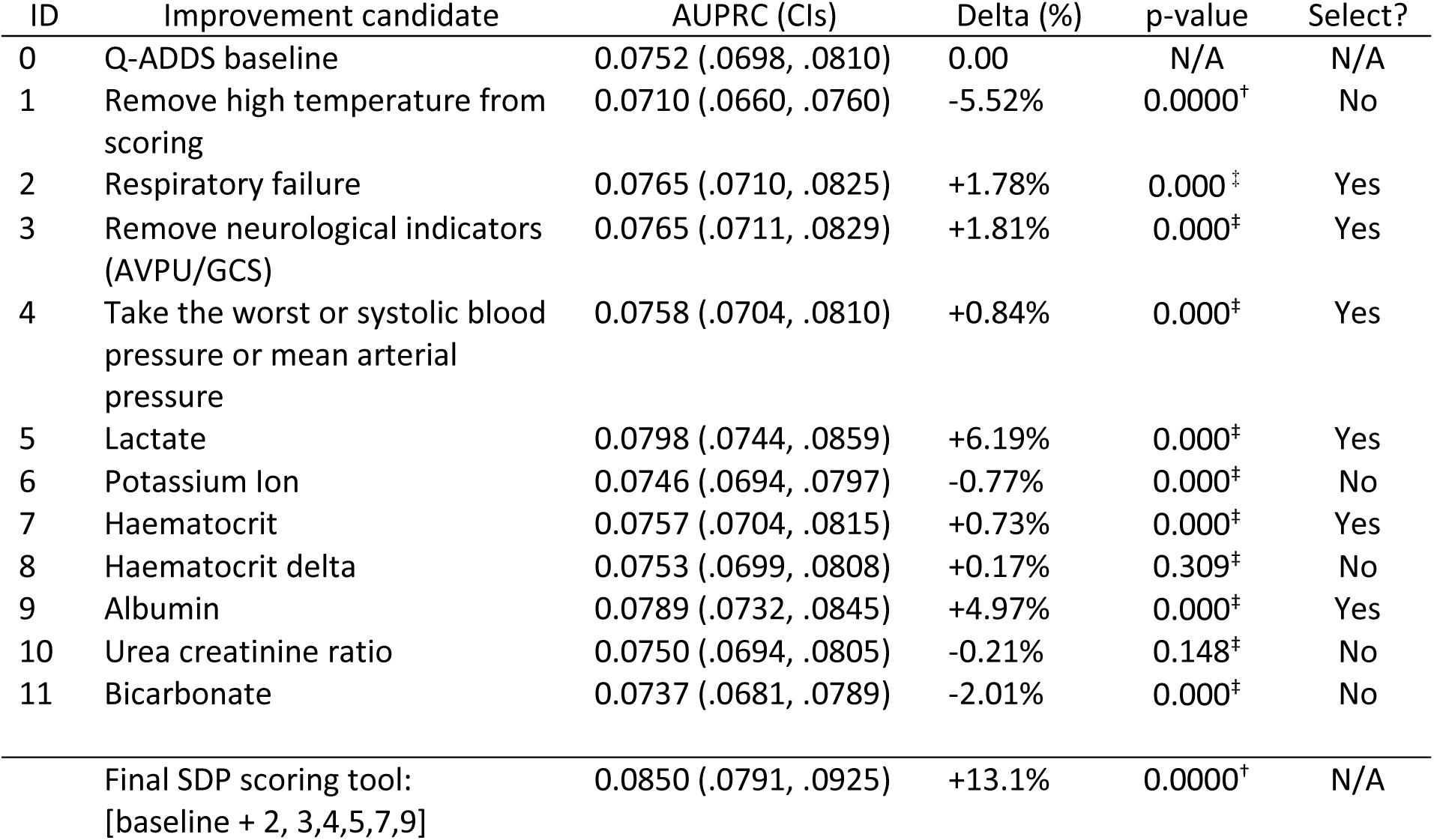
Area Under the Precision Recall Curve (AUPRC) results for the Q-ADDS baseline tool plus each improvement candidate (see **supplementary Table A2**) using the highest scoring data subset (5% of episodes) for each candidate. Also shown is the baseline tool and final SDP scoring tool, which includes the composite of the baseline and all Select=Yes improvement candidates. Delta is the percentage AUPRC difference between the augmented baseline plus candidate and the baseline. † Statistical difference assessed using Mann-Whitney U tests with Bonferroni correction ‡ Statistical difference assessed using ANOVA and Tukey HSD test (Levene’s test indicated no violation of homogeneity). Abbreviations, Q-ADDS = Queensland Adult Deterioration Score; AVPU = Alert Verbal Pain Unconscious; GCS = Glasgow coma score; AUPRC = Area under the precision recall curve; SDP = Seriously deteriorated patient.

**Table 3:** Final Seriously Deteriorated Patient (SDP) scoring tool variables together with their scoring ranges. Abbreviations, SpO_2_ = Peripheral Oxygen Saturation; O_2_ = oxygen; FiO_2_=Fraction of Inspired Oxygen; SBP = Systolic Blood Pressure; MAP = Mean Arterial Pressure.

| Variable/score | 4 | 3 | 2 | 1 | 0 | 1 | 2 | 3 | 4 |
| --- | --- | --- | --- | --- | --- | --- | --- | --- | --- |
| Respiratory rate | ≤8 |  |  | 9-12 | 13-20 | 21-24 | 25-30 |  | ≥30 |
| SpO<sub>2</sub> | ≤84 |  | 85-89 | 90-91 | ≥92 |  |  |  |  |
| Type 2 (hyper-capnic) Resp Failure |  |  |  |  |  |  |  |  | pCO<sub>2</sub> >65<br>AND<br>pH ≤ 7.3 |
| O<sub>2</sub> flow rate<br>Or<br>FiO<sub>2</sub> |  |  |  |  | ≤1 | 2-5 | 6-11 |  | >11 |
|  |  |  |  |  | ≤27 | 28-40 | 41-50 |  | >50 |
| SBP or MAP,<br>whichever score<br>is worse | <90 |  | 90-100 | 101-109 | ≥110 |  |  |  |  |
|  | <65 |  |  |  | ≥65 |  |  |  |  |
| Heart rate | <40 |  | 40-49 |  | 50-99 | 100-109 | 110-129 | 130-139 | ≥140 |
| Body temperature | ≤34 |  | 34.1-35 | 35.1-36 | 36.1 – 37.9 | 38-38.4 | ≥38.5 |  |  |
| Lactate |  |  |  |  | ≤2 |  | 2-3 | >3 |  |
| Hematocrit % |  |  |  |  | >39 |  | 20-39 | <20 |  |
| Albumin |  |  |  |  | >25 |  | 20-25 | <20 |  |

### Stage 3: Establishing the threshold for the Seriously Deteriorated Patient Indicator (SDPI)

The performance of the SDP scoring tool when evaluated between threshold settings of 10 and 13 is provided in **Table 4**. The clinician committee used this data and the selection criteria to decide on the final threshold: (i) *Superior accuracy for identifying unplanned ICU transfer patients than currently implemented MET thresholds*, where **Table 2** provides the superior AUPRC of the final SDP scoring tool (0.085) vs the Q-ADDS baseline (0.075); (ii) *Agreement that the score reflected seriously ill patients*, where at a threshold of 10, the SDP scoring tool was only marginally more precise (2-3%) than the baseline (Q-ADDS) MET threshold, so clinicians rejected this threshold as these patients may not be reliably ‘seriously’ deteriorated; (iii) *Similar or earlier identification of UPICU patients by SDPI relative to MET alert timing*, where an SDP scoring tool score of 12 and above were discarded because they identified UPICU patients 10-25% later than MET calls and were likely too restrictive, missing too many SDPs; and (iv) *A low chance that the SDPI threshold was reached due to a chronic organ failure or data entry errors alone*, where an SDP scoring tool score of 11 would require 3 or more severe value error entries or severe chronic organ failures. This left a score of 11 as the remaining alternative for the final threshold of the SDPI. This score also had higher odds ratio to both UPICU and death than the Q-ADDS baseline.

**Table 4:** SDP scoring tool performance and corresponding SDP cohort characteristics at different SDP scoring tool thresholds and for the baseline Q-ADDS MET level (score>=8) scoring tool. Abbreviations, Q-ADDS = Queensland Adult Deterioration Score; SDP = Seriously Deteriorated Patient; PPV = Positive Predictive Value; ICU = Intensive Care Unit; MET = Medical Emergency Team.

|  | Q-ADDs<br>@ MET | SDP scoring tool threshold scores |  |  |  |
| --- | --- | --- | --- | --- | --- |
|  |  | @10 | @11 | @12 | @13 |
| <u>Size of SDP cohort in patient episodes:</u> |  |  |  |  |  |
| Total | 18,179 | 18,532 | 12,323 | 8,313 | 5,532 |
| - Unplanned ICU transfers | 1,208 | 1,264 | 1,013 | 788 | 591 |
| - Died | 3,250 | 3,210 | 2,609 | 2,088 | 1,610 |
| - Recovered | 13,721 | 14,058 | 8,701 | 5,437 | 3,331 |
| <u>PPV: baseline result and % improvement over baseline for events:</u> |  |  |  |  |  |
| - Unplanned ICU transfers | 0.0665 | + 2.3% | + 24% | + 43% | + 61% |
| - Died | 0.1788 | - 3.1% | + 19% | + 41% | + 63% |
| <u>Median hours from first detection to event:</u> |  |  |  |  |  |
| Baseline result and % earlier detection of event over baseline for events: |  |  |  |  |  |
| - Unplanned ICU transfers | 6.7 | + 28% | + 7.5% | - 10% | - 25% |
| - Died | 63 | + 19% | + 13% | 0.0% | - 10% |
| <u>Odds Ratio</u> |  |  |  |  |  |
| Between index score and event: |  |  |  |  |  |
| - Unplanned ICU transfers | 23.3 | 24.4 | 27.6 | 30.1 | 32.6 |
| - Died | 42.9 | 40.9 | 46.8 | 53.5 | 60.8 |
| <u>Summary positions:</u> |  |  |  |  |  |
| Too imprecise; many patients not seriously deteriorated; susceptible to data entry errors | Yes | Yes |  |  |  |
| Too precise; likely missed many seriously deteriorated patients; late timestamp |  |  |  | Yes | Yes |
| Final score selected |  |  | Yes |  |  |

At a threshold score of 11, a seriously deteriorated outcome cohort of 12,323 (1.3% of total cohort) patient episodes are identified, of which 1,013 had an UPICU outcome (25% of UPICU outcome episodes, N=4,074), 2,609 died (33% of all deaths, N=7,997) and 8,701 (0.9% of cohort) had a recovery on the ward after a serious deterioration outcome, representing a 72% expansion (N=12,071 = UPICU + Death episodes) of the cohort with deterioration outcomes. Also, the SDPI outcome cohort were identified a median of 7.2 hours before UPICU and 71 hours before death.

## Discussion

The objective of this research was to develop a physiologically derived, replicable seriously deteriorated patient indicator (SDPI) that can be used as an outcome for evaluation of deterioration EWT performance as well as for training AI-DPAs. The proposed SDPI is derived from 13 commonly collected patient variables, including 7 vital signs, 5 laboratory variables and quantified oxygen requirement. The SDPI is rule-based (i.e., not AI derived) and therefore can be easily replicated by research groups who use standard datasets derived from their EMRs. It is designed to be used as a part of a composite outcome measure for potentially reversible deterioration, e.g., an outcome of UPICU or SDPI, whichever occurs first.

We correctly hypothesised that the SDPI would identify seriously deteriorating patients much earlier than the outcomes of UPICU or death: 7.2 and 71 hours earlier respectively. This is likely to have a considerable impact on EWT comparison and utility, for example, current EWTs typically target evaluation 24 hours prior to hospital death or UPICU.^8^ Our study has demonstrated that most deaths occur days after serious deterioration, so any preventive opportunity is missed with an alert that fires 48 hours after the deterioration has occurred, and EWT evaluation on this basis may be misleading for organisations seeking to implement them. This may also have a positive impact on AI-DPA training and subsequent accuracy, by focusing the training on the patient variable patterns leading up to serious deterioration, rather than after it when the patients may be undergoing palliative care prior to expected death.

We also correctly hypothesised that the SDPI would identify a significant cohort of seriously deteriorated patients that recovered on the ward who would otherwise be mislabelled as not deteriorated for evaluation and AI-DPA training purposes. In our dataset, the SDPI ‘recovery cohort’ consisted of 8,701 patient episodes, representing a 72% increase in the number of patient episodes with a deterioration event, as compared to UPICU and in hospital mortality alone. These episodes are likely to manifest as false-positives in most analyses, with corresponding adverse effects on AUPRC, AUROC, sensitivity, specificity, PPV and average accuracy. We acknowledge these effects would introduce a consistent bias in datasets, meaning it should be the same for all EWTs being evaluated, however reported EWT performance and rankings may differ significantly depending on each tool’s ability to predict the new ‘recovery cohort’. The SDPI also nearly doubles the outcome prevalence (when used in combination with UPICU and death); in our dataset, from 1.2% to 2.2%. Traditional deterioration outcomes have very low prevalence, causing underfitting during AI model development. This may be mitigated using over– or under-sampling techniques,^14,20,21^ but increasing the prevalence in a methodologically robust way is a superior solution.

Other research groups have done similar work. Byrd et al., developed an outcome on the basis of incipient trigger events indicating respiratory failure, bleeding and hypotension. ^22^ Their outcome triggers included mechanical ventilation and vasopressors, treatments that are not available in many ward environments. Similar limitations apply to the consensus-derived Adult Inpatient Decompensation Event.^23^ Research groups have also used MET or Rapid Response Team calls, which are system-based outcome measures with their own limitations including over-sensitivity and variations in hospital practices, protocols and documentation.^24–27^

### Strengths and Limitations

On a large, diverse multi-hospital dataset, we used a systematic, clinically grounded and physiologically plausible approach to develop a novel outcome measure that is needed to advance research on digital systems that predict clinical deterioration. ^6,22^ The SDPI reduces some of the mislabelling that has plagued the evaluation and development of both rule-based early warning scores and AI-DPAs. The SDPI provides more opportunity to identify patients with reversible clinical deterioration, and more correctly indicates the actual time that patients became seriously deteriorated. ^6^

Whilst we deliberately designed the SDPI to use commonly available variables and rule-based scoring ranges, we acknowledge others may prove superior and our future research involves a Delphi process to further refine the SDPI. Additionally, the method to derive the SDPI threshold of 11 was criteria and expert consensus based, which has limitations, however, the final SDPI threshold decision was also supported operationally by examining the odds ratio graph (**Supplementary Figure D2).** Until the OR of patients dying on the ward diverges from those transferring to ICU (point (a), SDP score ≤10), ICU-eligible patients are more likely to move to the ICU as their SDP scores increase. After the OR for UPICU patients plateaus (point (b), SDP score ≥13), as SDP scores increase, the likelihood of death also increases, indicating patients ineligible for ICU dying on the ward, whereas the likelihood of transferring to ICU plateaus and falls, indicating ICU-eligible patients have likely already transferred to the ICU. This supports scores of 11-12 being a ‘reversibility’ transition point.

The impact of variable data missingness on SDPI generalisability across different datasets requires further research. In our dataset, candidate variables such as potassium, bicarbonate, albumin and urea creatinine ratio were missing in around half of the episodes (see **Supplementary Table B3**). Despite this, the addition of many of these variables still improved the ability to identify seriously deteriorated patients. Had the missingness been lower, it is possible that more of these candidate variables may have been incorporated into the SDPI. Different evaluation datasets will have different missingness figures for each of these standard lab results and research is required to evaluate the impact of different variable missingness on the SDPI definition.

Finally, derivation of the SDPI was based on Australian patient data which may limit its external validity. Importantly, there are also patients who are deteriorated beyond what is reflected in the SDPI. This is because physiological decline can lag behind objective deterioration, and escalation due to clinician concern is often a better indicator of deterioration than any early warning score. ^28–30^

## Conclusion

This novel SDPI derived from EMR data provides a physiologically-derived, explainable and reproducible deterioration outcome measure that will support more accurate evaluation of EWTs by correctly identifying seriously deteriorated patients, and the time at which they deteriorate. While optimal ‘deteriorated’ and ‘deterioration’ definitions should continue to be sought, we propose the use of SDPI and UPICU for EWT evaluation and AI-DPA training.

## Acknowledgements

We acknowledge the ongoing support of Queensland Health, and specifically Clinical Excellence Queensland and the Queensland Digital Health Centre.

## Author Contributions

**Anton H. van der Vegt**: Conceptualisation, Methodology, Formal Analysis, Investigation, Data Curation, Writing – Original Draft, Funding Support; **Victoria Campbell**: Conceptualisation, Methodology, Investigation, Validation, Writing – Original Draft; **Imogen Mitchell**: Conceptualization, Validation, Writing – Review & Editing; **James Malycha:** Conceptualization, Validation, Writing – Review & Editing; **Ian A. Scott**: Conceptualization, Validation, Writing – Review & Editing; **Arthas Flabouris**: Conceptualization, Validation, Writing – Review & Editing; **Naitik Mehta**: Validation, Writing – Review & Editing; **Rudolf J. Schnetler**: Methodology, Formal Analysis, Investigation; **Christopher R Andersen**: Conceptualization, Validation, Writing – Review & Editing; **Daryl Jones**: Conceptualization, Validation, Writing – Review & Editing.

## Competing Interests

None declared

## Funding

AHV was funded through a Queensland Government, Advanced Queensland Industry Research Fellowship grant. The Queensland Government had no role within this research.

## Data availability

There is no data used in this study.

## Notes

### Competing Interest Statement

The authors have declared no competing interest.

### Author Declarations

This study was approved by the Townsville Hospital and Health Service Human Research Ethics Committee (HREC/QTHS/67897) and was granted a waiver of patient consent with Public Health Act approval

