## Supplementary Appendices for "Proposing a novel Seriously Deteriorated Patient Indicator (SDPI) for hospitalised ward patients"

#### Appendix A

##### Table A1: Strengths and weaknesses of existing clinical deterioration outcomes

| Outcome measure | Strengths | weaknesses |
|---|---|---|
| Death | • The most commonly used measure in this setting [1]<br>• Definitive outcome with no definitional variation<br>• Widely captured and consistent across all hospital locations | • The objective of early warning tools is not to predict the point at which death is imminent, but to predict preventable or reversible clinical deterioration<br>• Dying patients remaining on the ward after initial deterioration with death as an accepted and expected outcome may take many hours or days to die. During this interval intrusive laboratory tests are ceased, and the few, if any, vital signs that are taken can be profoundly deranged due to the dying process and/or symptom relieving medications. The resulting physiological profiles after onset of deterioration and before death is not the profile sought to identify deterioration in non-palliative patients<br>• Most (>90%) ward deaths in hospitals with mature rapid response systems are not unexpected[2] |
| Unplanned ICU transfer from the ward (UPICU) | • One of the most commonly used measure in this setting [1]<br>• Clinically valid as requires an intensive care specialist to accept patient for ICU level care<br>• Will capture the majority of 'unexpected' in hospital cardiac arrests (70% of in-hospital cardiac arrests receiving CPR who had no limitations on treatment gain return of spontaneous circulation (ROSC) and are transferred to ICU) [3] | • A late identifier of deterioration<br>• Many patients deteriorate but receive intervention and survive to discharge on the ward without ever getting to ICU or dying. If using only UPICU or death outcomes this cohort is mislabelled as never having deteriorated<br>• ICU resourcing and admission policies vary across jurisdictions, so that any one ICU admission may not be generalisable with respect to the admission threshold of severity of illness and reason for admission across a number of ICUs[4] |
| Cardiac Arrest | • Defined as having received CPR or defibrillation[5]<br>• Most accurately captures 'unexpected' deaths | • Rare in hospital wards where there are embedded, mature Rapid Response Systems; patients at risk are identified early such that the incidence of both predictable and preventable deaths is extremely low[3]<br>• Many cardiac arrests involve patients for whom appropriate treatment limitations have not been finalised and documented and therefore may include not-for-resuscitation patients<br>• This outcome makes the assumption that patients with treatment limitation orders (e.g., not for CPR) who die are 'expected' deaths. However, for most frail and comorbid patient admissions, while death in the next 12 months may not be a |

| | | |
|---|---|---|
| | | surprise, they are not expected to die during the current admission and most in fact survive to discharge. A death that occurs in these patients is actually a preventable death, but will be mislabelled as an ‘expected’ death<br>• Most patients in a non-critical care unit setting who achieve ROSC would go to ICU and therefore are identified within the UPICU outcome<br>• This outcome is not reliably documented in some EMRs. Often limited to retrospective datasets once ICD-10 coding of admission is available |
| Medical Emergency Team (MET) alerts/calls | • An earlier onset of non-critical deterioration<br>• A larger cohort with which to train algorithms<br>• Clinically valid as a clinician has activated the MET call | • The majority of MET calls/alerts do not involve patients who have seriously deteriorated, as evidenced by very few requiring critical care input or transfer to ICU ( <6% of patients with MET threshold breaches transfer to ICU in this dataset).[6,7] |

Abbreviations: ICU = Intensive Care Unit; UPICU = Unplanned transfer to the ICU; ROSC = Return of Spontaneous Circulation; CPR= Cardiopulmonary resuscitation; MET= Medical Emergency Team; ICD = International Classification of Diseases

#### Table A2: Candidate variables and rationale

| ID | Candidate to be tested | Baseline early warning score alteration | Candidate rationale |
|---|---|---|---|
| *1* | Remove high temperature from scoring | Body temperature > 36 = 0 | • High temperature is not an indicator of physiological failure<br>• Temperature can be influenced by provision of medications (e.g., paracetamol) |
| 2 | Type 2 Respiratory failure | pH level < 7.3 AND $pCO_2$ > 65 = 4 | • Indicator of true organ failure (ventilation failure)<br>• Requires blood gas and blood gas performance itself is an indicator of clinician concern for patient |
| 3 | Removal of low conscious level (AVPU/GCS) | AVPU/GCS <15 score = 0 | • Low conscious level is a common cause of MET escalation (~25%),[8] but is commonly due to transient events such as seizures or irreversible events, so most don't result in UPICU<br>• However, low conscious level is very commonly documented at the end of life<br>• Thus, low conscious level documentation may not be reflective of actionable, reversible deterioration |
| 4 | Low blood pressure (Take the worst score of either systolic blood pressure or mean arterial pressure) | Removed high SBP scores<br><br>lower SBP scores:<br>< 90 = 4; 90-100 = 2; 101-110 = 1<br>Add in MAP <65 = 4 | • High SPB alone unlikely cause for UPICU [9]<br>• Low SBP thresholds as per baseline Q-ADDS<br>• MAP included due to critical care inotrope decisions commonly based on MAP <65mmHg |
| 5 | Lactate | Between 2-3 = 2; > 3 = 3 | • Requires blood gas and blood gas performance itself is an indicator of clinician concern for patient<br>• Is increasingly performed with sepsis awareness campaigns<br>• Is scored as per sepsis definitions |
| 6 | Potassium Ion | 6 – 6.4 = 3<br>>6.4 = 4 | • Severe hyperkalemia is life threatening and requires monitored bed and aggressive treatment |
| 7 | Haematocrit | < 20 = 3<br>20-39 = 2 | • Scoring as per LAPS2 |
| 8 | Haematocrit delta (current – first recorded)/ first recorded) | >0.2 = 2 | • Fall may be more important than absolute value |
| 9 | Albumin | 20 – 25 = 2<br><20 = 3 | • Scoring as per LAPS2<br>• Low albumin is common in critically ill patients, and sepsis |
| 10 | Urea:Creatinine ratio | 40-80 = 1<br>>80 = 2 | • Scoring as per LAPS2 |
| 11 | bicarbonate | < 10 = 4<br>10-20 = 2 | • Acidosis in critical care is associated with increased mortality [10] |

**Table A2**: Candidate physiological scoring variables including the specific modification (column 3) applied to the baseline tool that was selected in Stage 1 and the rationale for the candidate. All other standard Vital signs (RR, $SpO_2$, Oxygen requirement, HR) scored as per baseline Q-ADDS. Abbreviations: Q-ADDS = Queensland Adult Deterioration Score; AVPU = Alert Verbal Pain Unconscious; GCS = Glasgow coma score; SpO2 = Peripheral Oxygen Saturation; O2 = oxygen; FiO2=Fraction of Inspired Oxygen; SBP = Systolic Blood Pressure; MAP = Mean Arterial Pressure, LAPS-2 = Laboratory-based Acute Physiology Score Version 2.0, MET = Medical Emergency Team, UPICU = unplanned Intensive Care admission.

### Appendix B: Data set exclusions

#### B1: Dataset exclusions: Method

During the period of data collection, a state-wide Cerner Millenium Electronic Medical Record (EMR) was being rolled out to eight of the eleven sites, with the other 3 sites having completed EMR implementation prior to the study start date. For the new EMR sites, their first 1000 patient admissions were excluded to avoid invalid data entries made during EMR implementation. Other patient exclusions included admissions devoid of at least one complete vital set, patients < 18 years of age, cross-hospital transfers within the same admission and non-inpatient admissions. We also excluded data pertaining to episodes of care outside of wards within highly monitored areas comprising the emergency department (ED), intensive care unit (ICU), operating theatres (OT) or post-anaesthetic care units (PACU)

#### B2: Dataset exclusions: Results

The original dataset comprised 1,480,148 admissions of which 648,423 were excluded because they were non-ward admissions (n=174,052), or had no minimum vital sign sets (n=474,371), the latter cohort being day-patients with a median length of stay of 5.2 hrs (IQR 3 – 6 hrs). This left 831,725 admissions for analysis, comprising 957,445 ward episodes involving 343,471 patients and 70,128,956 complete data samples (see **Table 1**).

#### Table B3: Dataset missing value counts

| | Missing % |
|---|---|
| ENCNTR_ID | - |
| Episode_num | - |
| avpu_or_gcs | 18.19 |
| avpu_val | 19.30 |
| fio2 | 88.12 |
| glasgow_cs | 47.92 |
| o2_flow_rate | 23.76 |
| rr | 1.62 |
| sp02 | - |
| urea_creat_ratio | 46.42 |
| albumin | 48.94 |
| haematocrit | 44.45 |
| age | - |
| mean_art_pres_calc | 6.64 |
| potassium | 47.10 |
| bicarbonate | 46.50 |
| hr_all | - |
| sbp_all | - |
| bt_all | 3.63 |
| lact_all | 83.69 |
| ph_all | 83.09 |
| pco2_all | 83.67 |
| po2_all | 83.72 |
| fio2_all | 65.59 |

### Appendix C: Early warning tool definitions

#### C1: National Early Warning Score (NEWS)

Introduced in the UK as paper EWT (NEWS) 2012, NEWS2 2017, digital implementation 2017, NEWS and NEWS2 are now used globally.
EWT type: Aggregate EW score
Scoring thresholds:

| Physiological Parameters | Score | | | | | | |
|---|---|---|---|---|---|---|---|
| | 3 | 2 | 1 | 0 | 1 | 2 | 3 |
| Respiration Rate (per minute) | ≤8 | | 9-11 | 12-20 | | 21-24 | ≥25 |
| SpO2 (%) | ≤91 | 92-93 | 94-95 | ≥96 | | | |
| Air or oxygen | | oxygen | | air | | | |
| Systolic Blood Pressure (mmHg) | ≤90 | 91-100 | 101-110 | 111-219 | | | ≥220 |
| Pulse (per minute) | ≤40 | | 41-50 | 51-90 | 91-110 | 110-130 | ≥131 |
| consciousness | | | | A | | | VPU |
| Temperature (ºC) | ≤35.5 | | 35.1-36.0 | 36.1-38.0 | 38.1-39.0 | ≥39.1 | |

Please note that AVPU and GCS results were combined as per table C4 below.

#### C2: Queensland Adult Deterioration Detection Systems (Q-ADDS) version 9.0

Introduced in Queensland, Australia as paper EWT 2011, digital implementation 2016
EWT type: Hybrid: Aggregate EW score + MET single trigger thresholds
Scoring thresholds:

Table B2: Revised Scoring table for Q-ADDS. Purple zone single trigger zones removed, only scoring component used, max score 4, to maintain scoring linearity

| EW Score | 4 | 2 | 1 | 0 | 1 | 2 | 3 | 4 |
|---|---|---|---|---|---|---|---|---|
| Respiratory Rate (brpm) | ≤8 | | 9-12 | 13-20 | 21-24 | 25-30 | | ≥30 |
| SpO2 (%) | ≤84 | 85-89 | 90-91 | ≥92 | | | | |
| Oxygen Flow rate or FiO2 (%) | | | | ≤1 | 2-5 | 6-11 | | >11 |
| | | | | ≤27 | 28-40 | 41-50 | | >50 |
| SBP (mmHg) | <90 | 90-100 | 101-109 | 110-159 | 160-169 | 170-199 | | ≥200 |
| Heart Rate (bpm) | <40 | | 40-49 | | 50-99 | 100-109 | 110-129 | 130-139 |
| Temp dCel | ≤34 | 34.1-35 | 35.1-36 | 36.1-37.9 | 38-38.4 | ≥38.5 | | |
| AVPU | PU | | V | A | | | | |

Please note that AVPU and GCS results were combined as per table C4 below.

#### C3: LAPS-2

Please refer to Escobar et al.[11] supplementary data, Table 3D.
Please note that AVPU and GCS results were combined as per table C4 below.

#### Table C4: Conversion between Glasgow Comma Score and AVPU

| GCS | AVPU |
|---|---|
| 14-15 | A |
| 9-13 | V |
| 4-8 | P |
| <4 | U |

Table C3: Conversion between Glasgow Coma Score (GCS) and AVPU (Alert, Verbal, Pain, Unconscious)

### Appendix D: Results

#### Table D1: Stage 1 results

| | NEWS index | Q-ADDS index | LAPS-2 index |
|---|---|---|---|
| AUPRC (CIs) | 0.0650 (0.0597, 0.0707) | 0.0752 (0.0698,0.0810) | 0.0585 (0.0530,0.0652) |
| Difference with: | | | |
| - Q-ADDS index | -15.7%* | | |
| - LAPS-2 index | +10.0%* | +22.2%* | |

**Table D1**: Rule-based deterioration index Area Under the Precision Recall Curve (AUPRC) comparison results using highest scoring data subset (5% of episodes) for each index. CI = Confidence Interval.

* Statistically different with adjusted p-value=0.0000 using Mann-Whitney U tests with Bonferroni correction

#### Figure D1: Comparative graph of Q-ADDS baseline (blue) and final SDPI scoring tool

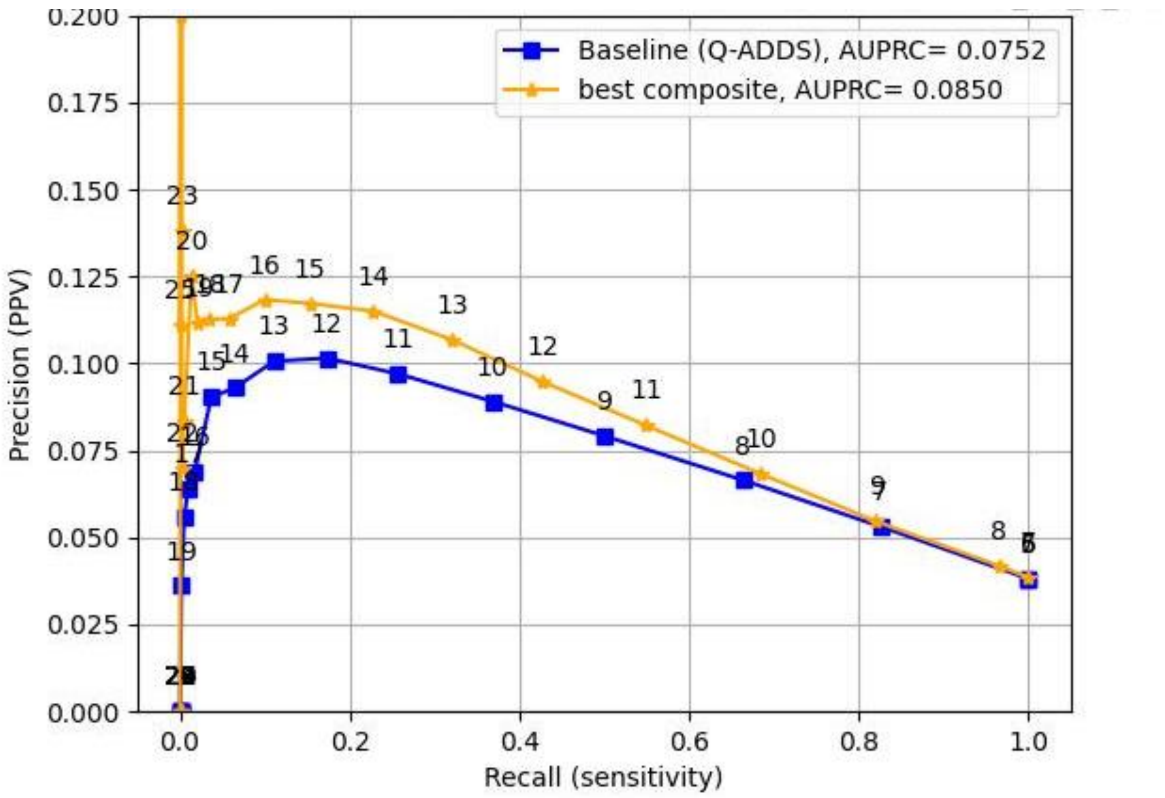

**Figure 3:** Precision-recall (PR) curves for the Q-ADDS baseline (blue) and final SDPI scoring tool (yellow, labelled composite) using the highest scoring data subset (5% of episodes) for each tool.

#### Figure D2: Odds ratio by SDP score for varied outcomes

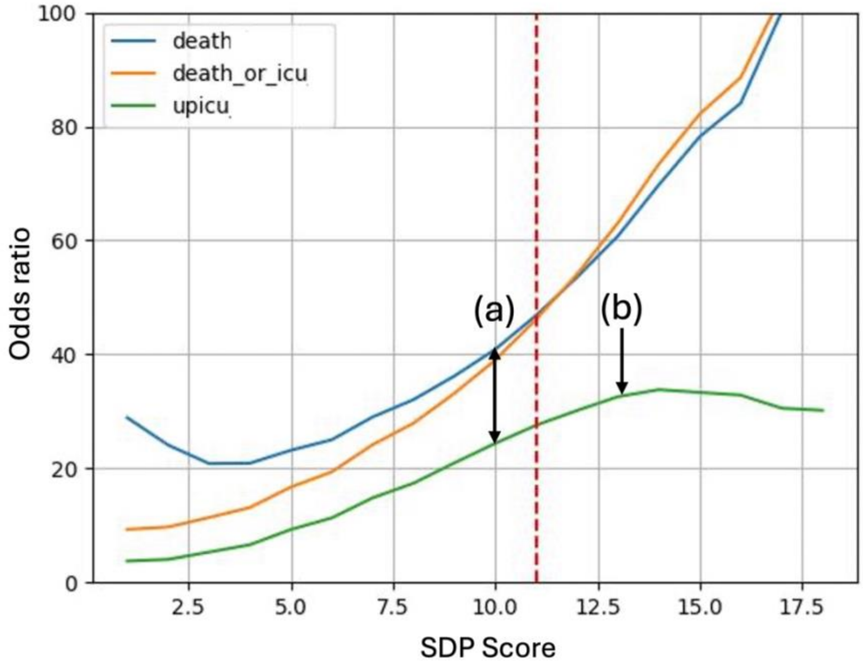

Figure D2: Odds ratio between each SDP score and the outcomes of death (blue), UPICU (green) or both (orange) and the point where the odds ratios for death and UPICU begins to diverge, (a), which approximates an SDPI score of 10, and ICU transfer begins to plateau, (b), which approximates a score of 13. Abbreviation, UPICU= unplanned transfer to the ICU; ICU = Intensive Care Unit, SDP = Seriously deteriorated patient.

11. Escobar GJ, Gardner MN, Greene JD, Draper D, Kipnis P. Risk-adjusting Hospital Mortality Using a Comprehensive Electronic Record in an Integrated Health Care

Delivery System. *Med Care*. 2013;51(5):446-453. doi:10.1097/MLR.0b013e3182881c8e
